# Evaluation of the evolution of SARS-CoV-2 Omicron variant and the spreading of LP.8.1 and NB.1.8.1

**DOI:** 10.1101/2025.06.16.25329694

**Authors:** Buqing Yi

## Abstract

Till now our understanding about the trend of evolution and spreading of SARS-CoV-2 is still limited. The fast spreading of newly emerging Omicron variants LP.8.1 and NB.1.8.1 has brought many questions regarding the evolution course and spreading trend. In this study we analyzed the evolutionary relationship among representative Omicron subvariants, and investigated spreading trend by analyzing the relative growth advantage of the newly emerging Omicron variants LP.8.1 and NB.1.8.1. The results have revealed a modest relative growth advantage of LP.8.1, and one much larger relative growth advantage of NB.1.8.1 over co-circulating BA.2.86/JN.1 subvariants including LP.8.1, indicating that NB.1.8.1 will possibly become the next dominant variant worldwide. As NB.1.8.1 is a product of intricate evolutionary events, the spreading of this variant would further complicate the prediction about future evolution and spreading trend of the SARS-CoV-2, highlighting the importance and necessity of performing genomic surveillance and monitoring SARS-CoV-2 evolution.

## 1. Introduction

It is important to predict the trend and impact of the evolution of SARS-CoV-2 regarding the public health implications. To acquire a better understanding about this critical topic, the evolution of SARS-CoV-2 has been under close monitoring globally. The newly emerging Omicron subvariants LP.8.1 and NB.1.8.1 have been spreading quickly worldwide, emphasizing the necessity to evaluate the newest evolutionary events of the Omicron variant and investigate the spreading of these variants.

The Omicron variant in general showed decreased hospitalization rates and less severe disease in patients [1], in comparison with the pre-Omicron SARS-CoV-2 variants, such as the Alpha or Delta variant. As one Variant of Concern (VOC), the Omicron variant has been circulating since late 2021, and many Omicron subvariants emerged, such as XBB.1.5, BA.2.86, JN.1, KP.3.1.1 and XEC [2, 3], and some of them have been separately identified as VOC or Variant of Interest (VOI). However, it is unknown if the new Omicron subvariants may further reduce pathogenicity.

In view of the significant growth globally and genetic changes, LP.8.1 was designated as a Variant under Monitoring (VUM) by the World Health Organization (WHO) in January 2025. Another Omicron subvariant NB.1.8.1 was detected a few months later than LP.8.1, and is growing rapidly compared to co-circulating variants. Similarly, on account of the mutations carried by NB.1.8.1 and the fast growth worldwide, NB.1.8.1 was designated as a VUM by the WHO in May 2025. Considering the relevance to public health, in this paper, we would focus on the evolutionary course of the Omicron variant and the spreading of the newly emerging variants LP.8.1 & NB.1.8.1 to acquire information regarding virus evolution and spreading, which might be able to help predict coming evolution and spreading trend of SARS-CoV-2.

## 2. Methods

### 2.1 Genome sequence and lineage classification

We used full-length SARS-CoV-2 sequences from GISAID [4] to build up genome sequence data set for investigation. We performed quality check and filtered out low-quality sequences that met any of the following criteria: 1) sequences with less than 90% genome coverage; 2) genomes with too many private mutations (defined as having >24 mutations relative to the closest sequence in the reference tree); 3) genomes with more than ten ambiguous bases; and 4) genomes with mutation clusters, defined as 6 or more private differences within a 100-nucleotide window. These are the standard quality assessment parameters utilized in NextClade (https://clades.nextstrain.org) [5]. We used the dynamic lineage classification method through the Phylogenetic Assignment of Named Global Outbreak Lineages (PANGOLIN) software suite (https://github.com/hCoV-2019/pangolin) [6].

### 2.2. Phylogenetic analyses of SARS-CoV-2

Phylogenetic analysis was carried out to investigate evolutionary relationship with a custom build of the SARS-CoV-2 NextStrain build (https://github.com/nextstrain/ncov) [7-10]. The pipeline includes several Python scripts that manage the analysis workflow. Briefly, it allows for the filtering of genomes, the alignment of genomes in NextClade (https://clades.nextstrain.org) [5], phylogenetic tree inference in IQ-Tree [11-13], tree dating [14, 15] and ancestral state construction and annotation. The phylogeny analysis is rooted by *Wuhan*-Hu-1/2019 (GISAID Accession ID: EPI_ISL_402125). Only samples fulfilling these criteria on GISAID were included in the analysis: 1. With complete sample collection dates; 2. With a complete sequence (>29,000nt) and less than 5% Ns.

The SRAS-CoV-2 VOCs/VOIs/VUMs BA.1, BA.2, BA.4, BA.5, BA.2.86, JN.1, XEC, LP.8.1 and NB.1.8.1 are included in the analysis. For each variant, ten samples (five from the early period and five from the late period of its circulation) were randomly chosen and used for the analysis.

### 2.3 Relative growth advantage

We analyzed relative growth advantage with SARS-CoV-2 genome sequences that were uploaded to GISAID with complete sample collection dates from the defined time duration and location. A logistic regression model was used to estimate the relative growth advantage of certain variant compared to co-circulating variants as previously reported [16-19]. The model assumes that the increase or decrease of the proportion of a variant follows a logistic function, which is fit to the data by optimizing the maximum likelihood to obtain the logistic growth rate in units per day. Based on that, an estimate of the growth advantage per generation is obtained (assuming the growth advantage arising from a combination of intrinsic transmission advantage, immune evasion, and a prolonged infectious period [20], and the relative growth advantage per week (in percentage; 0% means equal growth) is reported. The relative growth advantage estimate reflects the advantage compared to co-circulating variants in the selected region and time frame. The analyses were primarily performed with RStudio v1.3.1093 with multiple R software, e.g. tidyverse, ggplot [21-24].

## 3. Results

### 3.1. Mutation and recombination drive the evolution of the Omicron variant

Mutation is one of the major driving forces for evolution [25, 26]. In Table 1, the spike mutations detected in representative Omicron subvariants are listed. The evolution of the Omicron variant mainly took place on the basis of BA.2. The evolution from BA.2 to BA.4&BA.5 proceeded stepwise, with only a few novel spike mutations occurring in the background of the earlier lineage. It is noteworthy that XBB.1.5, as a product of recombination [27], owns about 10 novel spike mutations (some of them are also present in other XBB lineages). Furthermore, XBB.1.5 carries many mutations present in other VOCs/VOIs [28], such as V143- and Y144- from the Alpha variant [29]. Only a few mutations being frequently present in other VOCs/VOIs are absent in XBB.1.5, e.g. H69-; V70-; L452R [30].

**Table 1.** Spike mutation comparison among representative Omicron subvariants.

| Omicron BA.1 | Omicron BA.2 | Omicron BA.4&5 | Omicron XBB.1.5 | BA.2.86 | JN.1 | XEC | LP.8.1 | NB.1.8.1 |
| --- | --- | --- | --- | --- | --- | --- | --- | --- |
|  | T19I | T19I | T19I | T19I | T19I | T19I | T19I | T19I |
|  |  |  |  | R21T | R21T | R21T | R21T | R21T |
|  |  |  |  |  |  | T22N |  | T22N |
|  | L24- | L24- | L24- | L24- | L24- | L24- | L24- | L24- |
|  | P25- | P25- | P25- | P25- | P25- | P25- | P25- | P25- |
|  | P26- | P26- | P26- | P26- | P26- | P26- | P26- | P26- |
|  | A27S | A27S | A27S | A27S | A27S | A27S | A27S | A27S |
|  |  |  |  |  |  |  | S31- |  |
|  |  |  |  | S50L | S50L | S50L | S50L | S50L |
|  |  |  |  |  |  | F59S |  | F59S |
| A67V |  |  |  |  |  |  |  |  |
| H69- |  | H69- |  | H69- | H69- | H69- | H69- | H69- |
| V70- |  | V70- |  | V70- | V70- | V70- | V70- | V70- |
|  |  |  | V83A |  |  |  |  |  |
| T95I |  |  |  |  |  |  |  |  |
|  |  |  |  | V127F | V127F | V127F | V127F | V127F |
| G142- | G142D | G142D | G142D | G142D | G142D | G142D | G142D | G142D |
| V143- |  |  |  |  |  |  |  |  |
| Y144- |  |  | Y144- | Y144- | Y144- | Y144- | Y144- | Y144- |
| Y145D |  |  |  |  |  |  |  |  |
|  |  |  | H146Q |  |  |  |  |  |
|  |  |  |  | F157S | F157S | F157S | F157S | F157S |
|  |  |  |  | R158G | R158G | R158G | R158G | R158G |
|  |  |  | Q183E |  |  |  |  |  |
|  |  |  |  |  |  |  |  | G184S |
|  |  |  |  |  |  |  | F186L |  |
|  |  |  |  |  |  |  | R190S |  |
| N211- |  |  |  | N211- | N211- | N211- | N211- | N211- |
| L212I |  |  |  | L212I | L212I | L212I | L212I | L212I |
|  | V213G | V213G | V213E | V213G | V213G | V213G | V213G | V213G |
|  |  |  |  | L216F | L216F | L216F | L216F | L216F |
|  |  |  |  | H245N | H245N | H245N | H245N | H245N |
|  |  |  | G252V |  |  |  |  |  |
|  |  |  |  | A264D | A264D | A264D | A264D | A264D |
|  |  |  |  | I332V | I332V | I332V | I332V | I332V |
| G339D | G339D | G339D | G339H | G339H | G339H | G339H | G339H | G339H |
|  |  |  | R346T |  |  |  | R346T |  |
|  |  |  |  | K356T | K356T | K356T | K356T | K356T |
|  |  |  | L368I |  |  |  |  |  |
| S371L | S371F | S371F | S371F | S371F | S371F | S371F | S371F | S371F |
| S373P | S373P | S373P | S373P | S373P | S373P | S373P | S373P | S373P |
| S375F | S375F | S375F | S375F | S375F | S375F | S375F | S375F | S375F |
|  | T376A | T376A | T376A | T376A | T376A | T376A | T376A | T376A |
|  |  |  |  | R403K | R403K | R403K | R403K | R403K |
|  | D405N | D405N | D405N | D405N | D405N | D405N | D405N | D405N |
|  | R408S | R408S | R408S | R408S | R408S | R408S | R408S | R408S |
| K417N | K417N | K417N | K417N | K417N | K417N | K417N | K417N | K417N |
|  |  |  |  |  |  |  |  | A435S |
| N440K | N440K | N440K | N440K | N440K | N440K | N440K | N440K | N440K |
|  |  |  | V445P | V445H | V445H | V445H | V445R | V445H |
| G446S |  |  | G446S | G446S | G446S | G446S | G446S | G446S |
|  |  |  |  | N450D | N450D | N450D | N450D | N450D |
|  |  | L452R |  | L452W | L452W | L452W | L452W | L452W |
|  |  |  |  |  | L455S | L455S | L455S | L455S |
|  |  |  |  |  |  | F456L | F456L | F456L |
|  |  |  | N460K | N460K | N460K | N460K | N460K | N460N |
| S477N | S477N | S477N | S477N | S477N | S477N | S477N | S477N | S477N |
| T478K | T478K | T478K | T478K | T478K | T478K | T478K | T478K | T478I |
|  |  |  |  | N481K | N481K | N481K | N481K | N481K |
|  |  |  |  | V483- | V483- | V483- | V483- | V483- |
| E484A | E484A | E484A | E484A | E484K | E484K | E484K | E484K | E484K |
|  |  | F486V | F486P | F486P | F486P | F486P | F486P | F486P |
|  |  |  | F490S |  |  |  |  |  |
| Q493R | Q493R |  |  |  |  | Q493E | Q493E | Q493E |
| G496S |  |  |  |  |  |  |  |  |
| Q498R | Q498R | Q498R | Q498R | Q498R | Q498R | Q498R | Q498R | Q498R |
| N501Y | N501Y | N501Y | N501Y | N501Y | N501Y | N501Y | N501Y | N501Y |
| Y505H | Y505H | Y505H | Y505H | Y505H | Y505H | Y505H | Y505H | Y505H |
| T547K |  |  |  |  |  |  |  |  |
|  |  |  |  | E554K | E554K | E554K | E554K | E554K |
|  |  |  |  | A570V | A570V | A570V | A570V | A570V |
| D614G | D614G | D614G | D614G | D614G | D614G | D614G | D614G | D614G |
|  |  |  |  | P621S | P621S | P621S | P621S | P621S |
| H655Y | H655Y | H655Y | H655Y | H655Y | H655Y | H655Y | H655Y | H655Y |
| N679K | N679K | N679K | N679K | N679K | N679K | N679K | N679K | N679K |
| P681H | P681H | P681H | P681H | P681R | P681R | P681R | P681R | P681R |
| N764K | N764K | N764K | N764K | N764K | N764K | N764K | N764K | N764K |
| D796Y | D796Y | D796Y | D796Y | D796Y | D796Y | D796Y | D796Y | D796Y |
| N856K |  |  |  |  |  |  |  |  |
|  |  |  |  | S939F | S939F | S939F | S939F | S939F |
| Q954H | Q954H | Q954H | Q954H | Q954H | Q954H | Q954H | Q954H | Q954H |
| N969K | N969K | N969K | N969K | N969K | N969K | N969K | N969K | N969K |
| L981F |  |  |  |  |  |  |  |  |
|  |  |  |  |  |  |  | K1086R |  |
|  |  |  |  |  |  | V1104L | V1104L |  |
|  |  |  |  | P1143L | P1143L | P1143L | P1143L | P1143L |

- The direction of “left to right” indicates the timeline of Omicron subvariant occurrence with each variant on the left occurring earlier than the variant on its right, based on the time of earliest detection. - Yellow color indicates a novel mutation in a new position; Blue color indicates a novel mutation in a known active position. Here “novel mutations” are evaluated by comparing with pre-Omicron subvariants and previously occurred Omicron VOIs/VOCs/VUMs along with the time line, and the early Omicron subvariants BA.1, BA.2 and BA.4&BA.5 act as reference here. - The earliest detected B variant Wuhan-Hu-1/2019 (GISAID Accession ID: EPI\_ISL\_402125) serves as reference for mutation detection.

The Omicron variant went through one evolution jump that led to the appearance of BA.2.86, which carries around 20 novel spike mutations compared to the previous SARS-CoV-2 VOCs/VOIs. BA.2.86 was not derived from a further evolution of XBB.1.5 because none of the novel mutations in XBB.1.5* (in positions without previously detected mutation) was present in BA.2.86. The variant BA.2.86 further evolved to JN.1, which only has one additional mutation L455S in the background of BA.2.86. However, this single mutation significantly enhanced the fitness of JN.1 mainly by increasing immune evasion [31, 32], making JN.1 and its sub-lineages the predominant variants worldwide. Among the JN.1 subvariants, XEC is one product of recombination of two JN.1 sub-lineages KS.1.1 and KP.3.3 [33].

Novel mutations have two types: A. In known active sites (the positions where mutations have been detected in other previously identified VOCs/VOIs/VUMs) but novel mutations. B. In positions where mutations have been rarely detected previously. Compared to previously defined VOCs/VOIs/VUMs, LP.8.1 has one Type A novel mutation: V445R (V445P and V445H were previously detected in other variants); three Type B novel mutations: F186L, R190S, K1096R (some of them are also present in other LP lineages). The recently emerged variant NB.1.8.1 was first detected several months later than LP.8.1, but none of the LP.8.1 novel mutations occurs in NB.1.8.1. In comparison with previously defined VOCs/VOIs/VUMs, NB.1.8.1 also has both two types of novel mutations: one Type A mutation T478I (T478K was previously detected in other variants); two Type B mutations: G184S, A435S.

### 3.2. NB.1.8.1 shares a common ancestor with BA.2.86, but not a descendent of BA.2.86

We focus on phylogenetic distances among representative Omicron variants, in particular the VOCs/VOIs/VUMs that got spread globally, by “divergence” based on the relevant algorithm applied in Nextstrain. In Figure 1, the values of divergence of each variant from the earliest detected B variant are displayed. For B variant itself, the value of divergence is zero, indicating no divergence. The early Omicron variants BA.1 and BA.2 have a divergence of around 60. With the occurrence of BA.4 and BA.5, the divergence increased to around 80. Following a special speed-up evolutionary event, for the BA.2.86*/JN.1* lineages, the divergence escalated more than 50% compared to that of BA5. Previously the Omicron variants with a high divergence from the pre-Omicron variants showed pathogenicity changes producing milder symptoms [34], but till now there is no evidence demonstrating pathogenicity changes associated with BA.2.86/JN.1* lineages compared to the early Omicron variants.

**Figure 1.**
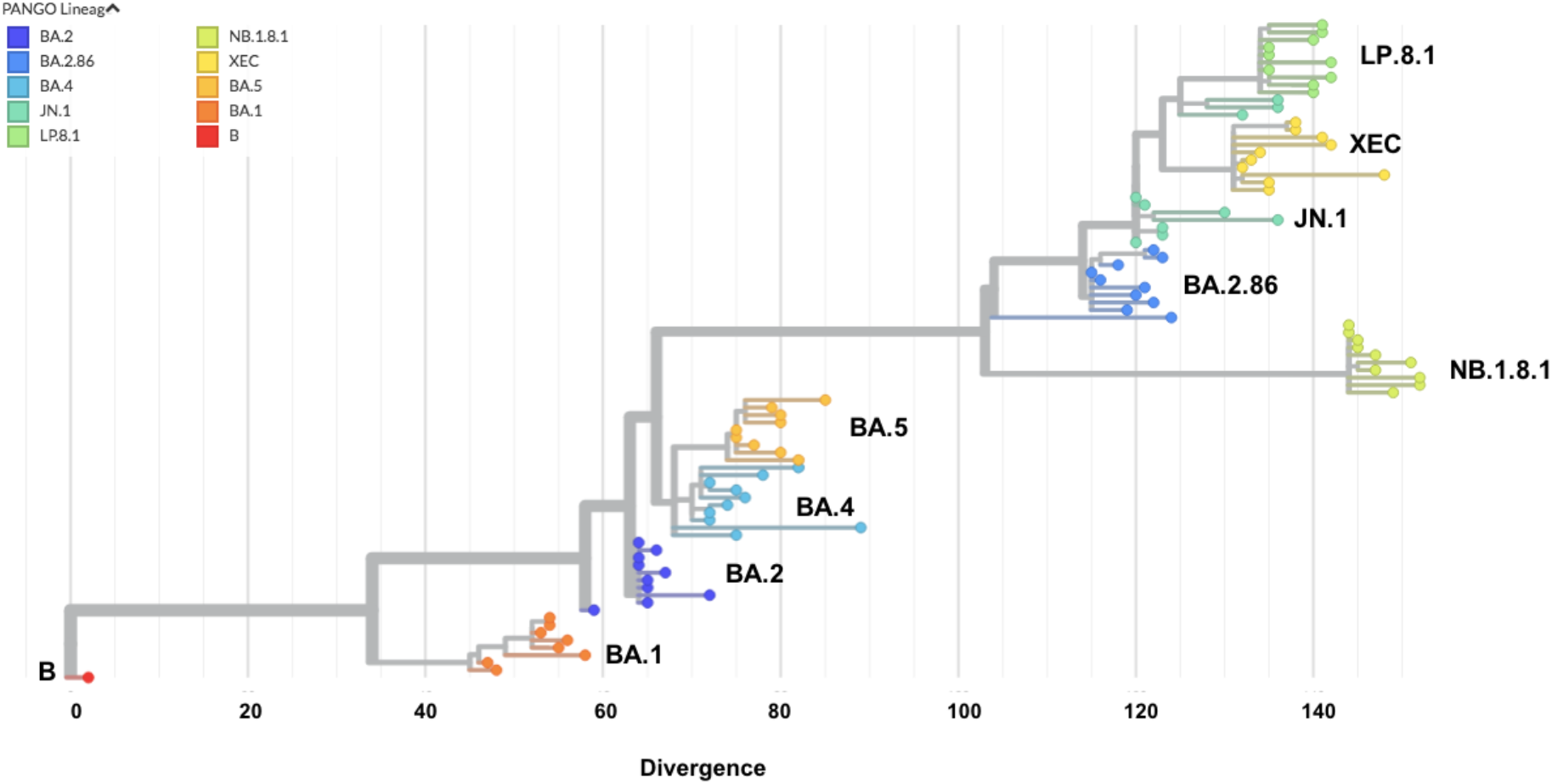
Evolutionary relationship between the representative Omicron VOCs/VOIs/VUMs. Each color represents one specific lineage. The names of VOCs/VOIs/VUMs are displayed next to the corresponding lineage groups. Numbers indicate phylogenetic distance of “divergence” calculated through the algorithm in Nextstrain. B is the earliest detected SARS-CoV-2 variant as reference for evaluating phylogenetic distance. For each Omicron subvariant, ten randomly chosen samples are used in the analysis.

Among the JN.1 sub-lineages, XEC was spreading quickly globally outcompeting many other JN.1 subvariants, but the divergence of XEC only slightly further increased compared to BA.2.86/JN.1*, showing a limited evolution from BA.2.86/JN.1*. Similar to XEC, LP.8.1 is another JN.1 subvariant that spread globally, the divergence of which also only slightly increased compared to BA.2.86/JN.1*, indicating a regular evolution process driven by mutations. It is noteworthy that from the evolutionary relationship, NB.1.8.1 shares a common ancestor with BA.2.86, but NB.1.8.1 is not one descendent of BA.2.86/JN.1, because a few BA.2.86 non-spike mutants such as N: Q229K and ORF1a: A211D, are missing in NB.1.8.1. This is consistent with genetic evidence that NB.1.8.1 is a descendent of XDV, a lineage derived from multiple recombination events either between different XBB sub-lineages or between one XBB sub-lineage and JN.1 [35, 36]. But there is also possibility that BA.2.86/JN.1* and XDV/NB.1.8.1* are the products of parallel evolution/convergent evolution/recombination from a common ancestor.

### 3.3. Global spreading of LP.8.1 and NB.1.8.1 and relative growth advantage

LP.8.1 was first detected in the USA on 2024-09-25, and till end of October 2024 this variant was reported from North America, Europe, Australia, and Asia based on available sequence data from GISAID. NB.1.8.1 was first detected in the Singapore on 2025-02-05. Till early April 2025, it was similarly reported from Asia, North America, Australia and Europe. In May 2025, globally the infection cases caused by LP.8.1 and its sub-lineages took up around one quarter of the total cases, and the infections caused by NB.1.8.1 and its sub-lineages also accounted for around 20% total cases. The fast spreading of these two variants suggests a relative growth advantage over co-circulating variants. Therefore, we analyzed the relative growth advantage of these two variants during there early spreading periods.

The relative growth advantage indicates the advantage over co-circulating variants in the selected region and time frame. As it has been reported that large scale community transmission often started or became detectable around one month later after the first sample was detected [2], to analyze the growth advantage during the early spreading period, we focus on the time period of approximately 45 days right after community transmission started. Considering the sequencing efforts are roughly comparable between 2024 and 2025, the results of relative growth advantage can be reliably compared to each other.

Globally, during the early spreading period (between 2024-11-01 and 2024-12-15), LP.8.1 showed a slight relative growth advantage of 11% over co-circulating variants, mainly XEC, KP.3.1.1 and a few other JN.1 subvariants (Figure 2A). In addition to evaluating the relative growth advantage taking all cases globally into consideration, we also specifically evaluated the relative growth advantage in North America. In North America LP.8.1 (between 2024-11-01 and 2024-12-15) showed a relative growth advantage of 24% (Figure 2A). The results indicate that LP.8.1 only displayed a modest relative growth advantage over co-circulating variants, e.g. XEC and KP.3.1.1, during the early spreading period.

**Figure 2.**
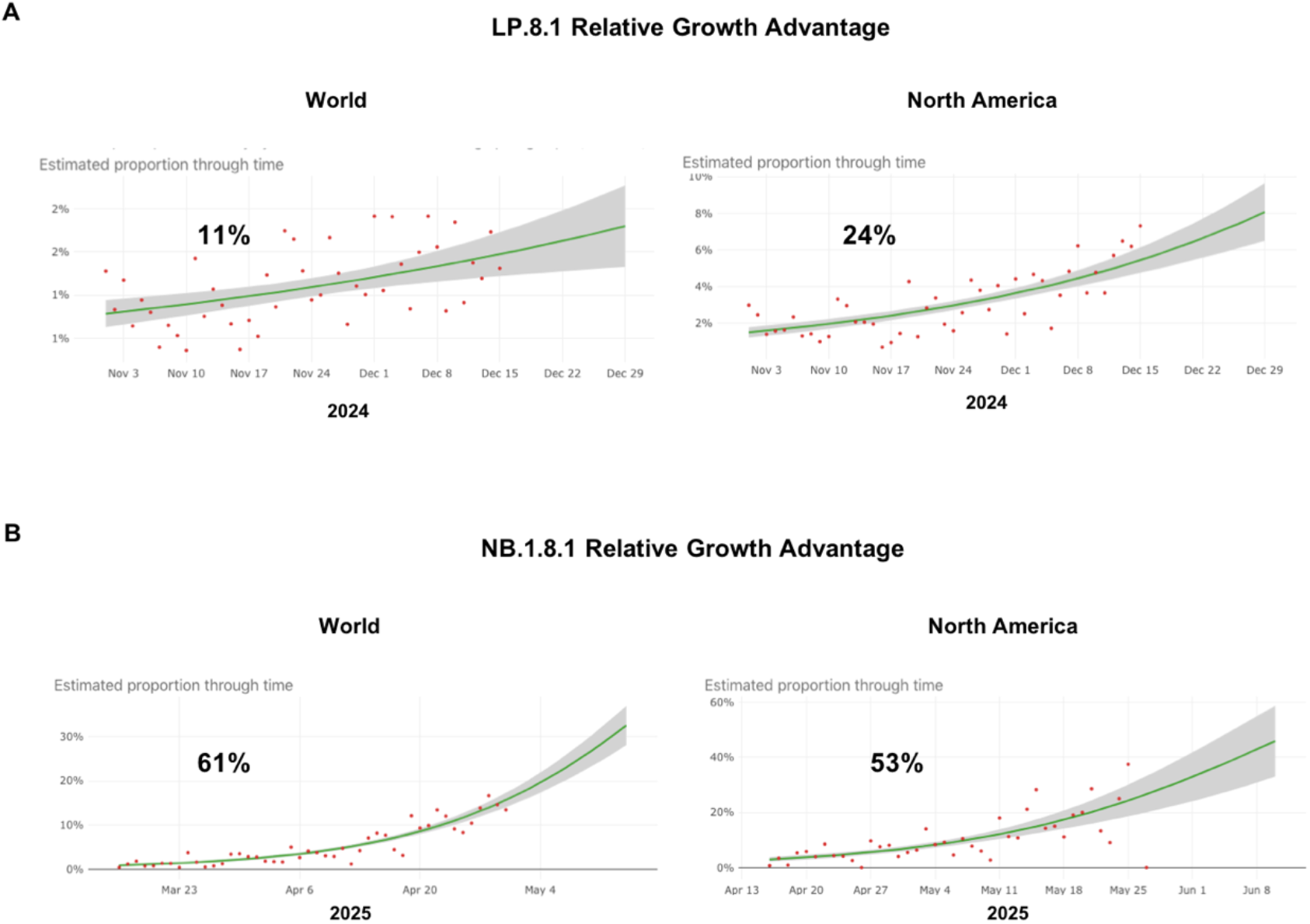
Growth of LP.8.1 and NB.1.8.1 during their early spreading period. Model fits are based on a logistic regression. Dots represent the daily proportions of variants. The relative growth advantage per week (in percentage; 0% means equal growth) is reported. The shaded areas correspond to the 95% CIs of the model estimates. A. Relative growth advantage of LP.8.1 in the whole world (Left) or in North America (Right). B. Relative growth advantage of NB.1.8.1 in the whole world (Left) or in North America (Right).

In contrast, in the whole world, during the early spreading period (between 2025-03-16 and 2025-04-30), NB.1.8.1 displayed a remarkable growth advantage of 61% against co-circulating variants, including LP.8.1 and multiple other JN.1 sub-lineages (Figure 2B). In North America, NB.1.8.1 (between 2025-04-16 and 2025-05-31) also displayed a high relative growth advantage of 53% (Figure 2B). This means, NB.1.8.1 has a growth advantage over LP.8.1, XEC, KP.3.1.1 and many other co-circulating Omicron subvariants, and possibly would become the dominant lineage worldwide.

## 4. Discussion

In this study, we evaluated the evolutionary relationship of representative Omicron variants, and investigated spreading trend by analyzing the relative growth advantage of the newly emerging VUMs LP.8.1 and NB.1.8.1. The results revealed a remarkable growth advantage of NB.1.8.1 over co-circulating BA.2.86/JN.1 subvariants, suggesting that NB.1.8.1 will possibly become the next dominant variant worldwide. Since NB.1.8.1 has a special genetic background as a product of recombination of multiple previous Omicron subvariants, the spreading of NB.1.8.1 would further complicate the prediction about future evolution trend of the SARS-CoV-2.

Following the global spread of JN.1, the subvariants of JN.1, such as KP.3.1.1 and XEC, emerged and rapidly spread worldwide. Since late 2024, LP.8.1, as one JN.1 subvariant, has also become one of the predominant variants in many countries worldwide. In this study, one moderate relative growth advantage of LP.8.1 over other co-circulating JN.1 subvariants during its early spreading period has been revealed. Consistent with that, V445R, a novel spike protein mutant on LP.8.1, has been shown to increase binding to human ACE2 receptors [37].

During the spreading of JN.1 subvariants, the impact of the XBB lineage, which was the dominant SARS-CoV-2 lineage in 2023 and early 2024 with XBB.1.5 & XBB.1.6 as the representative variants, has become more and more invisible. It is surprising that NB.1.8.1, one Omicron subvariant derived from the XBB lineage, occurred recently and rapidly spread. Despite its XBB background, NB.1.8.1 actually shares the spike protein with JN.1, with a few additional mutations [35]. In this study, we could show that NB.1.8.1 has a significant relative growth advantage over co-circulating SARS-CoV-2 variants, mainly JN.1 subvariants including LP.8.1. This is in line with the results from a few other virological investigations. It has been reported that the infection ability of NB.1.8.1 was higher than that of LP.8.1 [35], and the receptor binding domain of NB.1.8.1 could bind more strongly to human ACE2 receptor than that of LP.8.1 [38]. However, no difference was detected among NB.1.8.1, LP.8.1, XEC for the immune evasion ability targeting immunity triggered by XEC infection or JN.1 mRNA vaccine [35]. Virological studies in general indicate that NB.1.8.1 displays a balanced profile of ACE2 binding and immune evasion, suggesting a potential for future prevalence.

From evolutionary perspective, the changes taking place in NB.1.8.1 might have brough certain novel evolution opportunities, such as owing to the possible structural changes of NB.1.8.1 compared to other SARS-CoV-2 variants [39]. It complicated the prediction of future evolution trend, emphasizing the necessity of close monitoring of virus evolution.

In light of high levels of population immunity in many settings [40], in general the emergence of new SARS-CoV-2 variants would not pose a high risk on the public health. However, as we could not rule out the possibility of the occurrence of one variant that may lead to pathogenicity changes, such as due to co-infection or any special evolutionary event, it is necessary to monitor the virus evolution and acquire more comprehensive understanding about the evolution and spreading.

## Data Availability

All data produced in the present work are contained in the manuscript.

## Acknowledgements

We thank all researchers who are working around the clock to generate and share genome data on GISAID (http://www.gisaid.org). We thank the Dresden-concept Genome Centre for their sequencing efforts.

## Code availability

Data processing and visualization was performed using publicly available software, primarily RStudio v1.3.1093. Code for constructing phylogenetic maximum likelihood (ML) and time trees is available at https://github.com/genomesurveillance/delta-variant-sublineage, which is modified from SARS-CoV-2-specific procedures github.com/nextstrain/ncov.

